# Task shifting healthcare services in the post-COVID world: A scoping review

**DOI:** 10.1101/2023.02.26.23286301

**Authors:** Shukanto Das, Liz Grant

**Affiliations:** Usher Institute of Population Health Sciences and Informatics, Global Health Academy, Centre for Population Health Sciences, University of Edinburgh, Edinburgh, United Kingdom

**Author notes:** Corresponding author (SD).

## Abstract

Task shifting (TS) redistributes services from specialised to less-qualified providers. Need for TS was intensified during COVID-19 pandemic. We locate evidences of TS across all health conditions to answer: (A) What role has TS played in services delivery since the onset of the pandemic? (B) How has the pandemic impacted strategies of TS globally? We searched five databases in October 2022: Medline, CINHAL Plus, Elsevier, Global Health and Google Scholar. 35 citations were selected. Data was analysed thematically and reported as per PRSIMA-ScR. We used WHO health systems framework and emergent themes to discuss findings. TS was observed in countries of all income-levels. 63% (n=22) articles discussed what impact TS had in COVID-19 care, mental healthcare, care for HIV, sexual and reproductive health, nutrition and rheumatoid diseases. Others (n=13) highlight how pandemic altered TS strategies in mental healthcare, HIV services, hypertension and diabetes and emergency services. Studies varied in reporting TS; majority using terms “task shifting”, followed by “task sharing”, “task shifting and sharing” and “task delegation”. TS affected every block of health system. TS to non-specialists and non-healthcare staff improved services. Modifying roles through training and collaboration strengthened workforce. TS diagnostics increased access to medicines and technologies. Strategic leadership was key. Research on financing TS during pandemics is required. Stakeholders generally accepted TS. Shifting staff between programs led to unintended service incapacities. Pandemic affected strategies of TS. Training, providing care, follow-ups and consultations went digital. Virtually-delivered interventions improved outcomes. Accessibility to digital technology presented barriers. COVID-19 modified health-seeking behaviour. Patients preferred teleconsultations and online-symptom checkers. Organisations altered operating procedures and patient-flow pathways and added precautions to protect staff. Risks of spreading COVID-19 prompted facilities to reconsider TS. TS improved outcomes by filling workforce gaps and increasing access. We recommend TS to improve services delivery during the pandemic and beyond.

## Background

Critical shortage of skilled human resources for healthcare (HRH) vulnerates health systems and services to fragmentation. Projections of global HRH deficits by 2035 range between 12.9–18 million [1,2] Changing demographics and disease patterns, burdens of non-communicable diseases and emerging infections demands a larger qualified workforce. Given the correlation of healthcare provider numbers to service access, countries must instate appropriate ratios of skilled HRH to population through workforce development.[3] Coronavirus disease (COVID-19) has impacted services worldwide and slowed progress towards sustainable development goal–3 of achieving ‘healthy lives and well-being for all’ [4,5]. Countries have diverted finances, infrastructure and HRH to care for SARS-CoV-2 cases. Although resource-constrained settings have been disproportionately impacted, the pandemic has also affected nations with richer densities of facilities and HRH [6]. Nine among ten countries report considerable disruptions in essential healthcare services; aggravating disease burdens compared to pre-COVID years [7]. For instance, prevalence of anxiety and depression increased by 25%, with young people and women hit the worst [8]. Increase in depressive disorders costed over 49 million disability-adjusted life years [9]. Mortality due to human immunodeficiency virus (HIV) infections, tuberculosis and malaria are modelled to shoot by 10%, 20% and 36% respectively over the next five years [10]. Innovating diagnosis and treatment, scaling tele-healthcare and implementing task shifting and task sharing (TS/S) can sustain services [11].

TS/S rationally reallocate services from specialised staff to workers with lesser training or qualifications [12]. Often used synonymously, TS/S have differences. Task shifting (TS) focuses on providers assuming new roles through task delegation from one hierarchy to other. Task sharing is not territorial and allows providers to expand duties and collaborate [13–15]. Both approaches remove bottlenecks in service processes in resource-limited settings by engaging existing HRH efficiently or creating new cadres suitable to perform tasks; thereby increasing care access [13,16]. Redelegation is achieved by defining responsibilities to be shifted or shared and subjecting new cadres to competency-based trainings [16]. TS/S has been used as pragmatic responses to low HRH and acute healthcare demands. Russian Feldshers [17], French *Officiers de Santé* [18] and Chinese barefoot doctors [19] are historical examples. TS/S has been used in surgery and obstetrics [20] and anaesthesia and ophthalmic procedures [21]. Extensive studies in context of HIV in Africa, led to the World Health Organization (WHO) guidelines to adopt, invest and expand on TS/S, enable policies and facilitate implementation through services reorganisation [16].

Reviews have analysed how TS/S can benefit health systems beyond the pandemic. For example, shifting psychotherapy to trained non-specialists can increase timely access to mental healthcare (MH) [22–24]. Integrating TS into hypertension, diabetes and obstructive lung diseases management models could preserve care continuum during future pandemics [25] Task sharing family planning services with community healthcare workers (CHWs) can lessen pregnancy-related mortality and child morbidity [26]. Human papillomavirus screening and vaccine education through CHWs could help fight cervical cancer [27].

While literature on TS in individual diseases or health issues based out of one or few comparable contexts exists, to our best knowledge our review is the first to study TS as a tool in its own right and evaluate its impact on WHO health systems building blocks [28] since the onset of the pandemic and how TS evolved under the strains of COVID-19 globally. We undertake this scoping review to explore two questions: (A) What role has TS played in healthcare services delivery (HSD) since the onset of COVID-19 pandemic? (B) How has the pandemic impacted strategies of TS globally?

## Methodology

Scoping reviews help identify evidences and highlight gaps in a research domain [29,30]. We used the *population, concept* and *context* framework [31]. Our study *population* comprised of all healthcare providers, caregivers and HRH. We investigated the concept of TS services, including healthcare promotion, screening, diagnostics, treatment and rehabilitation, from more to less specialised HRH; across all health conditions. *Our context* was global; since the onset of COVID-19 pandemic (November 2019). Search words specific to ‘task shifting’, ‘task sharing’, ‘health services’ and ‘COVID-19’ were run across five databases in October 2022: Medline, CINHAL Plus, Elsevier, Global Health and Google Scholar. 177 citations were imported onto Covidence software [32]. Citations were included as per the population, concept and contexts discussed above. Citations in English were included. Reviews, commentaries and theses were excluded. Citations relating to task switching in cognitive science and learning were excluded. We extracted the following information from every citation: country and setting, reasons for TS, services shifted, HRH involved in TS, training and guidelines supporting TS, outcome measures of TS, stakeholder perspectives, key findings and author recommendations. We analysed extracted data thematically [34], using a deductive and inductive hybrid approach [35]. Detailed search strategy and protocol is included as supplementary material S1.

## Results

35 citations were selected in the final study (Fig 1). PRSIMA-ScR guidelines for scoping reviews [33] were followed to report findings (S2). 63% citations from our search discussed what impact TS had in HSD since the pandemic across areas of COVID-19 care (n=14), MH (n=2), HIV services (n=3), sexual and reproductive health (n=1), nutrition (n=1) and rheumatoid diseases (n=1). The other 37% articles discuss how the pandemic impacted strategies of TS in MH (n=7), HIV services (n=3), chronic illnesses, including hypertension and diabetes (n=2), alternative care modalities (n=1) and emergency medical services (n=1). Our evidences were from 25 countries. We used World Bank data[36] to find its distribution as: high-income countries (n=9), upper middle-income countries (n=5), low-and middle-income countries (n=9) and low-income countries (n=2). We used mapchart.net to visualise this distribution on a global map (Fig 2). We discovered variations in keywords and in-text terms used to articulate the phenomenon of TS from specialised to less-qualified HRH. 25 studies used the words “task shifting” (71.43%), 6 articles used “task sharing” (20.0%), 3 articles used “task shifting and sharing” (5.71%) and 1 article used “task delegation”. S3 lists the evidences of TS we found in HSD, since the pandemic. S4 lists our findings on the impact pandemic had on TS strategies. We discuss these findings in detail hereafter using the WHO health systems framework [28] and other emergent themes.

**Fig 1:**
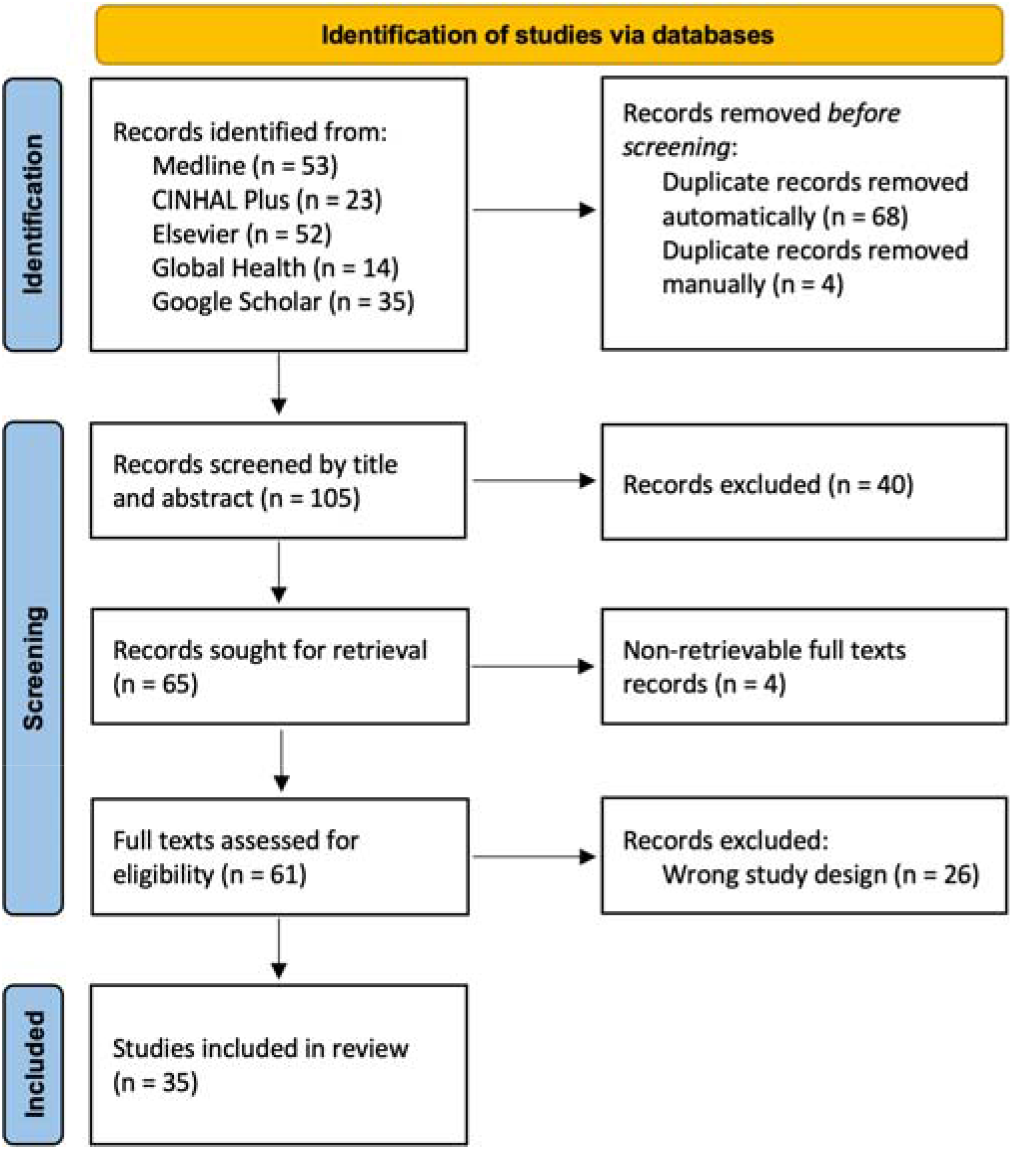
PRSIMA flow diagram of search and selection process of citations.

**Fig 2:**
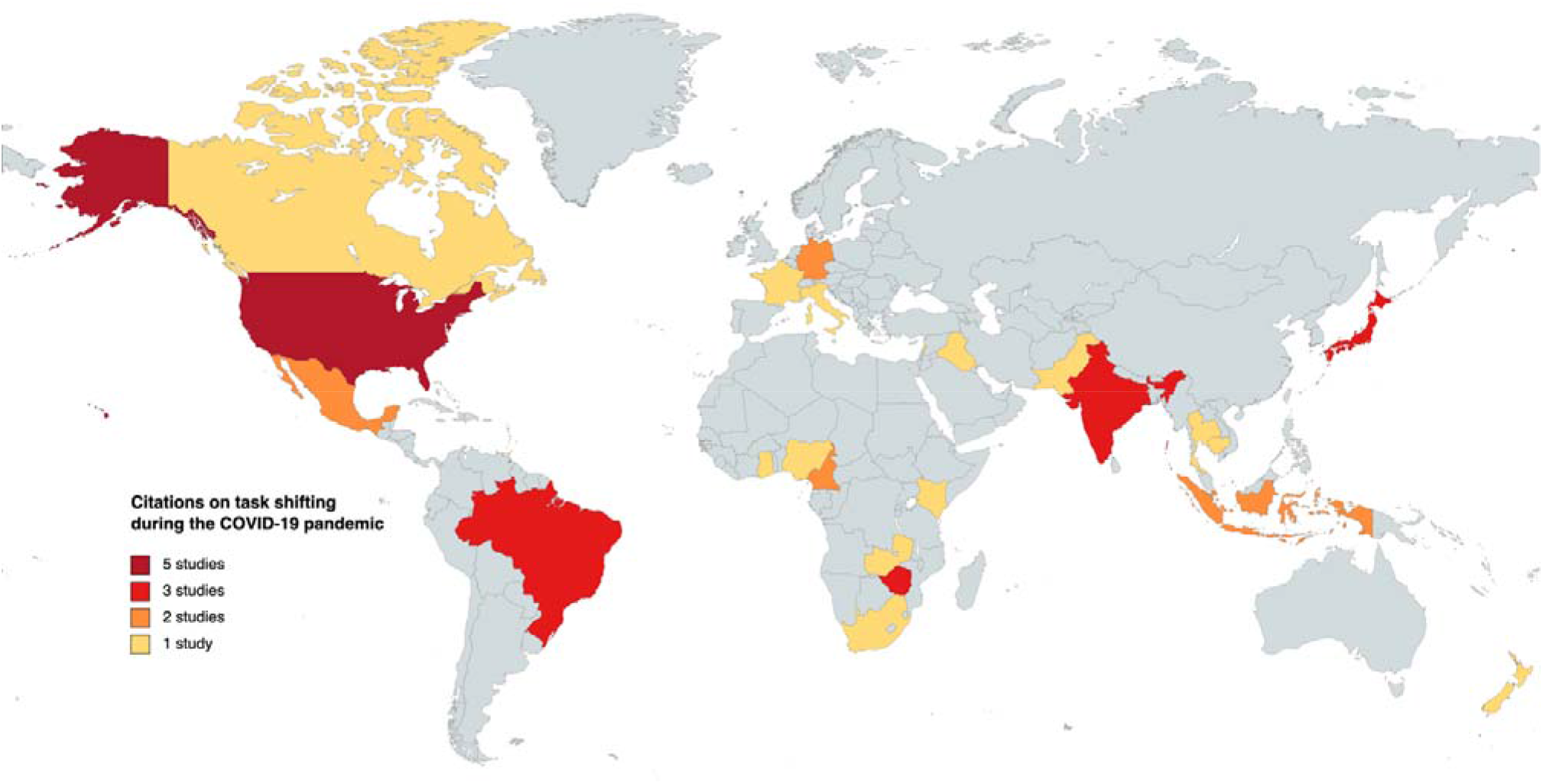
Global distribution of task shifting in healthcare services delivery since the onset of COVID-19 pandemic.

## Discussion

Reallocation of HRH and other resources towards pandemic response strengthened COVID-19 care, but impacted other HSD [37]. Our analyses of how TS was engaged in HSD and how the pandemic in-turn affected strategies of TS, is presented under the following two sections.

### (A) Role of TS in HSD since the pandemic

As early as in March 2020, the WHO recommended use of TS/S during the pandemic [38] In this section, we use the WHO health systems framework [28] to discuss how TS impacted its six interconnected building blocks; progress in each being necessary for achieving improved healthcare access, coverage, quality, safety and social and financial risk protection [39].

#### Service delivery

This unit entails to safe effective interventions offered to those in need when required, while minimising wastages [40]. With increasing SARS-CoV-2 cases, Governments prioritised epidemiological surveys for infections. Case study by Chidavaenzi *et al* [41] on a community-wide rapid antigen-based serial screening program for tribe members at detention centres and casinos in San Carlos Reservation demonstrated that TS testing to trained non-clinical staff catalysed reduction in infection transmission. Non-clinical staff from local health departments and staff at detention centres and casinos were upskilled in sample collection, reporting, confidentiality and administration. They performed 3,834 tests among 716 people over 28 days, whereby only one person turned positive. Although community immunity through previous infections, vaccinations and social distancing may have reduced cases, TS serial testing to non-clinical staff certainly lowered transmission [41]. Success of this program demonstrated that trained non-healthcare staff can aid in community screening programs.

COVID-19 widened MH needs and service gaps. Ortega *et al* [42] described Mexican non-profit Compañeros En Salud and the Chiapas health ministry building MH capacity through TS and skilling of non-specialist providers. Primary care physicians, CHW and non-clinical staff were trained in psychological first aid, referrals and self-care and were given pocket guides to extend psychosocial support to community members. With increasing SARS-CoV-2 cases, physicians and nurses from respiratory clinics were also skilled in psychosocial care and rehabilitation. CHW were trained in contact tracing and their home-visit questionnaires evaluated anxiety and suicidal thoughts. Furthermore, they provided MH to patients grieving and in need of palliative MH due to COVID-19. Depending on severity, providers referred to higher care [42]. TS expanded coverage of MH during the pandemic and incorporated instruments to address pandemic-induced stressors. This approach to MH, presents potential solutions to fill gaps in low-resource settings.

#### Health workforce

Adequate numbers of competent, compassionate and responsive HRH forms this building block [40]. During the pandemic, HRH responsibilities were moved outside their specialties to meet demands. For example, multi-country survey by Taylor *et al* [43] revealed that primary care physicians were reassigned to intensive care (USA) and final year medical students (Brazil) assisted with SARS-CoV-2 caseloads. Case study of tertiary hospital in Singapore by Davis *et al* [44] remarked that nutrition behaviour contracts for patients and caregivers, conventionally charted by psychologists, were drawn by physicians and nurses. Study of public health nurses (PHN) in Tokyo by Honda *et al* [45] reported nurses experiencing anxiety, frustration and fatigue due to sudden increase in workloads. PHN identified tasks, including contact tracing, health monitoring, hospitalisation coordination and consultation with residents, to be shifted onto part-time PHN and other nursing and support staff to forestall organisational dysfunction [45]. Similarly, analysis of teleconsultations by PHN during the first infection wave by Yoshioka-Maeda *et al* [46] showed that conversations were majorly about prevention measures and referral pathways, which PHN need not have attended. Managers were advised to let PHN handle essential consultations and shift teleconsultations onto low-level staff and office workers by developing script-based manuals and proper training. TS freed PHN to focus on infection control and management. Role modifications, however need to be complemented by changes in staff organisation and management. Abraham *et al* [47] described central teaching hospital in Ghana changing staff roles and rosters to complement transferring medication prescribing and dispensing antiretroviral therapy (ART) at pharmacies to nurses. TS ensured continuity of ART and reduce patient waiting time at HIV clinics.

While TS refers to transfers from more to less specialised workforces, the pandemic precipitated instances where more-qualified HRH assumed roles conventionally undertaken by less-qualified HRH. Taylor *et al* [43] reported primary care physicians undertaking fieldwork to screen, test and triage patients (Singapore, Trinidad and Tobago) and increase awareness about social distancing, symptoms and quarantining (Iraq, India). Shortage of nursing staff led primary care physicians to perform nursing procedures and therapy (Italy). Raskin *et al* [48] reported that since dental clinics, dental hygienists were pulled into community screening and telehealth services to educate community members. While countries differed on HRH utilisation strategies, TS through rational reorganisation, cross-training and collaboration provided stopgap solutions to manage COVID-19 across countries.

#### Health information systems

Reliable timely production, analysis, dissemination and usage of health system indicators constitutes this subunit [40]. Routine clinical audit data are evidences that drive quality improvement in hospitals [49] During the pandemic, sub-standard management of medical records led to system failures in clinical auditing. Multi-hospital based South African study by Sono-Setati *et al* [50] recommended TS routine clinical data collection and audit reporting from clinical staff, particularly from emergency healthcare providers to other HRH, to reinforce care and coordination. This will help reduce mortality, morbidity, stressors and burn outs. PHN at Japanese public health centers shifted teleconsultations onto office staff as discussed previously [46] Yoshioka-Maeda [51] reported PHN developing web-based data systems to exchange COVID-19 patient information and shift hospital coordination and clerical tasks to office staff, and inventory management, including management of personal protective to external companies. TS through web-based systems saved nursing time, decreased workloads and improved efficiency in allocating supplies.

#### Access to essential medicines and technologies

This unit entails to cost-effective use of high-quality products, including medicines, vaccines and other technologies [40]. Patent access to diagnostic tools was vital as SARS-CoV-2 cases increased. Centralised testing facilities, limited HRH and poor logistics added to Zimbabwean struggles in meeting demands, as per Gudza-Mugabe *et al* [52]. To increase access, build testing capacity and reduce reporting time, Zimbabwe followed WHO guidance of September 2021 [53] of TS antigen-based diagnostics onto laboratory technicians. Technicians were upskilled in testing and machines were upgraded with COVID-specific software. Decentralising testing from one to over thousand centres through TS, increased testing availability and uptake, shortened report turnaround time to under a day and curtailed staff burnouts [52].

### Financing

Subunit on financing encompasses mechanisms to fund healthcare and protect users from catastrophic expenditures [40]. Our search did not report economic evaluations of TS since the pandemic. TS saving costs for low-and middle-income countries, particularly in ART, is discussed elsewhere [54,55]. However, we found a study by Omam *et al* [56] discussing the success of Cameroonian differentiated service delivery model in decentralising ART to the community. Mobile HIV units were established through this model and testing, counselling, ART initiation and refilling and the tasks of following up and linking lost-to-follow-up patients with treatment were shifted onto non-clinical staff within primary care and community-based facilities. Although the differentiated service delivery model improved access to HIV services in hard-to-reach conflict regions, authors emphasise the need for economic evaluations of TS-based models before scaling across more conflict settings [56]

### Leadership and governance

This block pertains to effective stewardship in building accountable strategic policies and regulatory structures [40]. Timely preparedness plans and implementation aided pandemic management. Case study by Köppen *et al* [57] on Germen Federal and State policies on TS reported that while Federal polices recommended TS, TS was not incorporated by states. Federal infection control law authorised TS from doctors to paramedics, considering they held competencies, were not handling serious patients and performed documentation. However, it did not list these tasks or competencies. Only state policies of Saxony-Anhalt supplemented information to Federal law by mentioning competencies as per treatment protocols and enabling tools for documentation [57]. Although one state contextualised Federal guidelines successfully, lack of specificity and direction led to failures in adopting TS nationwide.

Foresighted direction at all organisational levels is key. Hospitals recognised the need for TS intensive care. Helmi *et al* [58] evaluated capacities of intensive care units (ICU) in Indonesia and reported inadequacies in rooms, equipment and specialists distribution. Given these limitations, leaderships directed TS in ICU through training on rights, responsibilities, communication, coordination and insurance schemes and incentives for involved HRH.[58] Mukhsam *et al* [59] described strategies by University Malaysia Sabah, Borneo, to mitigate impact of COVID-19 within campus, with a focus on MH through TS. Strict policies on screening, usage of personal protective equipment and attendance were implemented. Dedicated teams performed surveillance, health promotion, quarantining and sanitation. Upon return, international Chinese students were screened and quarantined for two weeks. Local mandarin-speaking medical students were tasked to supervise quarantined students on online-messaging applications and provide psychosocial support, health education and ensure they filled home-monitoring questionnaires. These measures resulted in no case importations into the campus [59]. Quick policy intervention followed by effective delegation and management helped attain zero-COVID status.

Policy makers must identify and respond to threats in order to safeguard interventions. Case study by Pry *et al* [60] on Zambian COVID-19 mitigation guidelines for HIV demonstrated that prompt policy and implementation reduced interruptions in ART delivery. The government recommended dispensing six multi-month ART to patients and TS patient communication and mobilisation onto trained lay providers. These guidelines led to greater than a four-fold increase in early collections of ART. Weekly receipt of multi-month dispensation increased from 47.9% to 73.4%, and proportion of late visits fell from 18.8% to 15.1% [60]. Policy based on TS by lay providers improved ART uptake during the pandemic.

#### Unintended consequences of TS

Alongside positive systemic impacts, TS produced certain consequences. Interviews of district managers and HRH in Islamabad by Zafar *et al* [61] reported staff undergoing reorganisation to meet acute needs of the pandemic. Government-employed lady health workers and dengue outreach workers were trained on surveillance and communication. These CHW were deployed into the community to identify SARS-CoV-2 cases and bring patients to testing. TS contact tracing to CHW however, caused them to abandon their commitments with other services, including maternal health and immunisation programs, and the sudden vacuum of HRH and service incapacity, caused these to suffer.

Faria de Moura Villela *et al* [62] assessed pandemic’s impact on Brazilian HRH and reported that three-quarters of their doctors, nurses and hospital staff experienced structural changes and role-shifts. COVID-19 wards, ICU and emergency departments witnessed highest TS/S. While this assisted in managing SARS-CoV-2 cases, role changes and corresponding salary cuts coincided with elevated anxiety and depression among staff. Pandemic changed doctor-patient relationships and nature of practice teams. Eggleton et al [63] reported receptionists at General Practitioners (GP) in New Zealand being upskilled in telephone triaging and routing patients to consultations, nursing care or other facilities. Nurses and receptionist teams ran separate respiratory units where they triaged patients with possible symptoms to prevent transmission. These were seen by GP separately. While TS improved outputs and freed GP to attend more patients, managing different patient streams added to workloads of nurses and receptions, necessitating support staff hires.

#### Acceptability of TS

Providers show resistance to new cadres taking on tasks. Consulting with stakeholders to help them appreciate rationale behind TS/S can motivate HRH to accept task redelegations [64,65]. Interviews by Jacobi *et al* [66] illustrated how success in contraceptives and medicines dispensing by CHW increased access to family planning services across humanitarian–development nexus and led stakeholders view TS necessary during workforce crunch. Likewise, Kuhlmann *et al* [67] describe how rheumatologist shortage in Germany led care for rheumatoid arthritis and other musculoskeletal disorders shift onto GP and rheumatology specialist assistants. Interestingly, rheumatologists found TS onto rheumatology specialist assistant more useful, compared to delegating onto GP. Exacerbated workforce shortages primed stakeholders towards TS; showcasing feasibility and increasing acceptability. Institutionalising TS can further service access beyond the pandemic.

Medical students contribute to bridging care-provision gaps. While their engagement in pandemic response has varied across nations [68], their clinical involvement to fill HRH shortages has been recommended [69]. Surveys by Mohammed *et al* [70] across teaching hospitals in Nigeria assessed student knowledge and provider-willingness for COVID care. 90% students expressed willingness to assist but remarked fears of infections and parental disapproval. Medical students are regarded as healthcare experts by their communities [70]. TS care onto them, complemented with suitable training, can be an effective acceptable approach to increase service capacity.

### (B) Role of COVID-19 on strategies of TS

Next, we review how the pandemic impacted implementation strategies and processes used in TS. Our analysis informed on two sub-themes:

#### Digital technology enabled capacity building

Through capacity building, organisations address healthcare needs by developing services, infrastructure, HRH and supporting structures [71]. COVID-based containment caused interventions cease routine activities and rapidly adopt new delivery models. SUMMIT trials by Singla *et al* [72] is TS behavioural activation for perinatal depression and anxiety from psychologists, psychiatrists and social workers onto nurses and midwives. Lockdowns in USA and Canada caused training and supervision of providers, follow-ups and data collection to migrate onto patient-centred virtual systems. PROACTIVE by Scazufca *et al* [73] examined effectiveness of psychoeducation-based support to older adults as delivered by trained CHW with no higher education or formal training in MH. Lockdowns in Brazil caused therapy to move onto telephones. Trial found 62.5% participants recover from depression, compared to 44% recovery in participants receiving usual care. Jordans *et al* [74] described that compared to training as usual, online-delivered competency-driven training improved facilitator ability in offering psychological treatment to distressed adolescents in Lebanon by 18% without increasing training duration. These trials underscore the potential telemedicine-supported community MH interventions have in improving service quality and access.

Two Indian studies on substance use and addiction demonstrate how digital training and monitoring improved MH screening and provider knowledge, attitude and practice. Nirisha *et al* [75] compared performance of accredited social health activists (AHSA) receiving in-classroom training, with ASHA receiving additional virtual training; latter actualised by COVID-19. Virtually trained ASHA identified significantly higher number cases of alcoholism (83%; p⍰≤⍰0.00). Their knowledge, attitude and practice scores improved more compared with in-classroom trained ASHA. In another study by Philip *et al* [76], primary care physicians were virtually trained and monitored by specialists. Post training, their knowledge scores improved by 37%. 80.7% and 47.7% trainees expressed confidence in identifying mental health issues in patients and caregivers respectively. 60.5% trainees felt confident in prescribing and managing patients, 64.9% in providing deaddiction services and 52.6% successfully identified when to refer. These cases show potential of scaling online-based MH build capacity at primary care level. Remote training offers sustainable strategies by improving time efficiency and reducing SARS-CoV-2 infection risks, travel costs and carbon emissions.[77,78] Accessibility and affordability of digital technology are limiting factors.

Friendship Bench, which shifts screening and psychological support for common mental disorders to trained grandmothers, switched to digital applications during lockdowns. Dambi *et al* [79] piloted chat-based application Inuka, and discovered its implementation in Zimbabwe as feasible and acceptable. Depression and anxiety of users declined and quality of life improved. Kamvura *et al* [80] further integrated Friendship Bench with screening for comorbid conditions, diabetes and hypertension; maximising on TS using a theory of change approach to reduce tensions between new and existing providers [65]. Digital platforms can catalyse TS in MH and non-communicable disease care. But connectivity, application instability, expensive mobile data and power outage can limit scaling up [79].

Same-day ART initiation to patients diagnosed with HIV increases ART uptake, viral load suppression and care retention [81]. Lujintanon *et al* [82] are decentralising ART initiation to trained lay providers in Bangkok. Due to COVID, monitoring and physician consultations are being conducted virtually. Lessons from this TS-based trial will help overcome geographical and HRH barriers [82].

COVID-19 changed how healthcare was sought, delivered and regulated online. Telehealth utilisation increased 38 times and investments, tripled; fuelling innovation to improve access and affordability [83]. French survey by Oikonomidi *et al* [84] gauged perceptions of ideal post-pandemic balance between traditional and alternative modalities, such as teleconsultations, remote monitoring and symptom checking, shifted from physicians onto applications. 22% patients with chronic illnesses preferred online symptom-checkers instead of contacting physicians when new symptoms appeared. Respondents considered TS symptom checking to online platforms as appropriate pre-consultation tools. These were found inappropriate for patients with anxiety, heterogeneous symptoms or symptoms only physicians could diagnose. Patients expressed that TS pre-consultations to applications need accreditation to quality control authorities and supervision by physicians [84].

#### Modifications in standard operating procedures

HRH shortage and reduced financial support [85] limit ART and pre-exposure prophylaxis (PrEP) delivery in Africa. Coulaud *et al* [86] reported district-level centres in Cameroon TS routine ART prescription and administration to drive better outcomes of ART uptake. In Kenya, one-stop shop (OSS) models improved PrEP delivery, uptake and continuation. Roche *et al* [87] described how relocating client files, equipment and drugs to OSS, and TS PrEP dispensing to lower cadres reduced provider movement and waiting times. However, social distancing necessitated testing points closest to OSS be shut. Clinics transformed OSS into isolation centres and moved OSS operations to community clinics farther away. OSS reorganisation meant HRH and patients travelled between centres; wasting time and productivity [87].

Organisations modified standard operating procedures to protect HRH through the pandemic. Compañeros En Salud trains non-specialists in MH as discussed previously [42]. They extended their scope to support pandemic-based stress and anxiety. As per Rodriguez-Cuevas *et al* [88], providers undertook regular patient-home visits outdoors and while visiting suspected cases, donned N95 or KN95 masks, face shields, isolation gowns and gloves.

Neonatal ICU in Cambodia shift incubator-side tube feeding and bag-and-mask resuscitation onto fathers and grandmothers. Iwamoto *et al* [89] report how risks of family transmitting SARS-CoV-2 to patients and staff, necessitated additional thermal scanning, sanitation and a second line of screening through symptom-based questionnaires and contact tracing. Authors further reported facilities seeking to hire additional nurses and train them to reduce dependence on task sharing with family caregivers [89]. This, however, would reverse progress made with TS.

#### Limitations and implications of future research

We evidenced that TS from specialised to less-qualified HRH increased efficiency of HSD through the pandemic. However, this was synthesised from published literature originating from a finite set of nations, which did not include many low-income countries. Several of our citations provided inadequate information on which tasks were transferred or which HRH were involved. Research evaluating how guidelines for TS are framed, implemented and feedbacked into by HRH involved, particularly during emergencies, is required. Economic evaluations of TS in light of pandemics is also needed.

## Data Availability

All relevant data supporting the findings are within the paper and its supporting information files.

## Conclusions

Our review discovered TS occurring through the pandemic in countries of all income-levels across areas of healthcare, including COVID-19 care, MH, HIV services and others. TS was necessitated due to acute healthcare demands, HRH shortages and incapacities caused by the pandemic. Multiple terminologies were used to refer to TS. TS affected every building block of health system. TS demonstrated that non-specialists and non-healthcare HRH reinforce community-based programs and offer sustainable solutions to fill workforce gaps. Modifying HRH roles, building reliable information systems, training and corporation strengthened workforces and expanded coverage and access. Strategic leadership at all levels was key in sustaining services through TS. Stakeholders accepted TS, given magnified demands and workforce crunch. TS HRH from one healthcare vertical to the other however, unintentionally led to service incapacities in donor programs. Through suitable training and monitoring, TS can improve HSD during and beyond the pandemic.

The pandemic affected mechanisms of TS considerably. Interventions delivered by non-specialists migrated onto patient-centered digital platforms. Decentralisation of HSD was supported through specialist consultations virtually. These transitions improved outcomes, particularly at primary care level. Remotely delivered interventions introduced safe time- and cost-effective solutions to HSD, but accessibility to digital technology presented challenges. COVID-19 altered health-seeking behavior of patients, who now prefer teleconsultations and online-symptom checkers. Quality assurance and supervision by experts can help increase faith in these aides. Standard operating procedures of healthcare facilities and programs changed, and service points and patient-flow pathways underwent reorganisation. Organisations added precautionary measures to protect staff. Risks of infection spread due to sharing tasks with external caregivers however, prompted facilities to consider hiring additional staff. This movement was counterproductive to TS.

## Author Contributions

Both authors contributed to the conceptualisation of the study. LG advised on the approach and methodology. SD developed the search protocol, analysed the data and drafted the manuscript. LG reviewed the data analysis and the manuscript. Authors received no specific funding for this work. Authors declare no conflict of interest.

## Supplementary information

**S1: Protocol used to conduct literature search and data analysis**

**S2: PRISMA-ScR checklist for reporting data**

**S3: Examples of task shifting in healthcare services delivery since COVID-19 pandemic**

**S4: Examples of the impact of pandemic on task shifting strategies**

