## Supplementary information 1 for "Task shifting healthcare services in the post-COVID world: A scoping review"

**Protocol for literature search and data analysis**

We conducted a scoping review as they help understand the range, scale and nature of a research theme, outline and present evidences, highlight gaps in literature and help evaluate whether undertaking a full systematic review pertaining to the question of interest would be beneficial [1,2]. Scoping reviews differ from systematic reviews in a way that the latter usually has well-defined questions and study designs to begin investigating literature with, while the former has a broader focus and can include diverse types of studies in it [2]. We follow the methodological framework of 5 stages [2] to conduct this scoping review:

**Stage 1: Identifying the research question**

Our aim has been to gather evidences from literature exploring two questions:

A. What role has task shifting played in healthcare services delivery since the onset of COVID-19 pandemic?
B. How has the COVID-19 pandemic impacted strategies of task shifting globally?

**Stage 2: Identifying relevant studies**

An initial search was carried out on Medline via PubMed on October 07, 2022 using PubMed. This search returned 136 articles. But on screening titles and abstracts, over 1/2 citations were found irrelevant hinting at the need of refinement of query code.

Following this, the search strategy was revised, the search code was rewritten and nw run across a total of 5 databases between October 11-12, 2022:

1. Medline via PubMed (<https://pubmed.ncbi.nlm.nih.gov>)
2. CINHAL Plus via EBSCO (<https://www.ebsco.com/products/research-databases/cinahl-database>)
3. Elsevier via Scopus (<https://www.scopus.com>)
4. Global Health via OVID (<https://ovidsp.ovid.com>)
5. Google Scholar (<https://scholar.google.com>)

**Search terms for Medline PubMed is shown here as sample:**

(("healthcare"[Text Word] OR "health"[Text Word] OR "healthcare programs"[Text Word] OR "healthcare delivery"[Text Word] OR "health care organisation and systems"[Text Word] OR "health systems"[Text Word] OR "delivery of health care"[Text Word] OR "health plan implementation"[Text Word] OR "healthcare workforce"[Text Word] OR "healthcare provider*"[Text Word] OR "healthcare provider training*"[Text Word] OR "caregiver"[Text Word] OR "health service*"[Text Word] OR "service delivery"[Text Word] OR "health services use"[Text Word] OR "primary care"[Text Word] OR "community clinic"[Text Word] OR "rural health clinics"[Text Word] OR "rural health services"[Text Word] OR "primary care management"[Text Word] OR "patient care management"[Text Word] OR "primary care services"[Text Word] OR "healthcare providers"[Text Word] OR "models of care"[Text Word] OR "clinical management"[Text Word] OR "emergency care"[Text Word] OR "preventive care"[Text Word] OR "preventive healthcare"[Text Word] OR "preventive medicine"[Text Word] OR "public health"[Text Word] OR "health promotion"[Text Word] OR "hospitalization"[Text Word] OR "hospital care"[Text Word] OR "surger*"[Text Word] OR "surgical care"[Text Word] OR "palliative care"[Text Word] OR "rehabilitative care"[Text Word] OR "curative care"[Text Word] OR "patient-centered care"[Text Word] OR "patient centred-approach"[Text Word] OR "patient-centred care"[Text Word] OR "person-centred approach"[Text Word] OR "person-centred care"[Text Word] OR "human resources for health"[Text Word] OR "patient care team"[Text Word] OR "healthcare teams"[Text Word] OR "community health worker"[Text Word] OR "community health worker training*"[Text Word] OR "community nurs*"[Text Word] OR "ayush"[Text Word] OR "ayurveda"[Text Word] OR "homeopathy"[Text Word] OR "asha"[Text Word] OR "anganwadi"[Text Word] OR "medical informatics"[Text Word] OR "traditional medicine"[Text Word] OR "barefoot doctor*"[Text Word] OR "physician assistant*"[Text Word] OR "physician associate*"[Text Word] OR "non-physician clinicians"[Text Word] OR "mid level health worker*"[Text Word] OR ("Health Care Category"[MeSH Terms] OR "Occupational Groups"[MeSH Terms] OR "Disciplines and Occupations Category"[MeSH Terms] OR "analytical, diagnostic and therapeutic techniques and equipment category"[MeSH Terms] OR "Diseases Category"[MeSH Terms] OR "Psychiatry and Psychology Category"[MeSH Terms]) OR ("health policy"[Text Word] OR "healthcare policy"[Text Word] OR "policy implementation"[Text Word] OR "healthcare strategy"[Text Word] OR "universal health coverage"[Text Word] OR "health systems research"[Text Word] OR ("Public Policy"[MeSH Terms] OR "Organizational Policy"[MeSH Terms] OR "Diseases Category"[MeSH Terms])) OR ("quality of healthcare"[Text Word] OR "Quality of Health Care"[Text Word] OR "quality of care"[Text Word] OR "healthcare outcomes"[Text Word] OR "patient satisfaction"[Text Word] OR "universal health coverage"[Text Word] OR "access to care"[Text Word] OR "evidence-based treatment"[Text Word] OR "performance measures"[Text Word] OR "healthcare evaluation"[Text Word] OR "quality of life"[Text Word] OR "health services research"[Text Word] OR "health workforce development"[Text Word] OR "health workers planning"[Text Word] OR "health human resources planning"[Text Word] OR ("Quality of Health Care"[MeSH Terms] OR "Diseases Category"[MeSH Terms])) OR ("human capital"[Text Word] OR "healthcare investments"[Text Word] OR "health economics"[Text Word] OR "sustainable healthcare"[Text Word] OR "cost-benefit analysis"[Text Word] OR ("Health Care Economics and Organizations"[MeSH Terms] OR "Diseases Category"[MeSH Terms]))) AND ("task shift*"[Text Word] OR "task shift*"[Text Word] OR "TS/S"[Text Word] OR "task shar*"[Text Word] OR "task shar*"[Text Word] OR "task shifting and task sharing"[Text Word] OR "task transfer*"[All Fields] OR "task delegat*"[Text Word]) AND ("2019-ncov disease"[Text Word] OR "2019-ncov disease"[Text Word] OR "2019-ncov diseases"[Text Word] OR "covid"[Text Word] OR "covid19"[Text Word] OR "covid 19"[Text Word] OR "sarscov"[Text Word] OR "sars cov 2"[Text Word] OR "corona virus"[Text Word] OR "coronavirus"[Text Word] OR "coronavirus diseases"[Text Word] OR "novel coronavirus"[Text Word] OR "covid pandemic"[Text Word] OR "covid19 pandemic"[Text Word] OR "covid-19 pandemic"[Text Word] OR "sars-cov2 detection"[Text Word] OR "sars-cov-2 diagnosis"[Text Word] OR "long covid"[Text Word] OR "long covid19"[Text Word] OR "long covid-19"[Text Word] OR "post covid"[Text Word] OR "post covid19"[Text Word] OR "post covid-19"[Text Word] OR "covid management"[Text Word] OR "covid treatment"[Text Word] OR "covid management"[Text Word] OR "covid-19 management"[Text Word] OR "covid-19 clinical management"[Text Word] OR "covid treatment"[Text Word] OR "covid19 treatment"[Text Word] OR "covid-19 treatment"[Text Word] OR (("COVID-19 Testing"[MeSH Terms] OR "COVID-19 Serological Testing"[MeSH Terms] OR "covid 19"[MeSH Terms] OR "COVID-19 Vaccines"[MeSH Terms] OR "sars cov 2"[MeSH Terms] OR "post-acute COVID-19 syndrome"[Supplementary Concept] OR "adult multisystem inflammatory disease covid 19 related"[Supplementary Concept]) AND "pediatric multisystem inflammatory disease covid 19 related"[Supplementary Concept]))) AND ((fha[Filter]) AND (fft[Filter]))

Results of the search is shown as under:

| **Search Query** | **Databases** | **Medline**  11 Oct, 2022 | **CINHAL** 11 Oct, 2022 | **Elsevier**  12 Oct, 2022 | **Global Health**  12 Oct, 2022 | **Google Scholar**  12 Oct, 2022 |
| --- | --- | --- | --- | --- | --- | --- |
| **Healthcare**  (Healthcare delivery OR Healthcare policy OR Quality of healthcare OR Health economics) | | 28,164,950 | 2,362,831 | 9,069,952 | 1,310,086 | About 2,540,000 |
| **COVID-19** | | 314,238 | 271,712 | 437,661 | 97,191 | About 638,000 |
| **Task Shifting OR Task Sharing** | | 2,316 | 976 | 4,116 | 798 | About 17,300 |
| **(Task Shifting OR Task Sharing) AND Healthcare** | | 2,183 | 826 | 2,108 | 785 | 153 |
| **(Task Shifting OR Task Sharing) AND Healthcare AND COVID-19** | | 53 | 23 | 52 | 14 | 35 |
| **Total unique citations after removing duplications** | | | | | | **105** |

**Stage 3: Selecting studies**

Citations were imported into Covidence systematic review management software (<https://app.covidence.org>). Duplications were removed automatically and manually. Citations were screened as per the following selection criteria. PRSIMA-ScR guidelines [3] were followed to screen and report citations (see Fig 1 in the paper).

| **Inclusion criteria** | **Exclusion criteria** |
| --- | --- |
| - All articles on task shifting services from higher to lower professional cadres of healthcare providers and caregivers - All geopolitical contexts and care settings - All diseases, health conditions and healthcare problems - Citations published since the onset of COVID-19 pandemic, i.e. November 2019 - Citations available in English language only | - Articles on delegation of work between same cadres of healthcare providers and caregivers - Studies relating to task switching in cognitive science and learning - All literature reviews, commentaries, editorials, dissertations and conference proceedings |

**Stage 4: Charting of the data**

We read full-texts and extracted the following information from the identified citations independently:

- Title of the paper
- Author information
- Journal of publication
- Date of publication
- Aim of the study
- Focus on task shifting and/or task sharing
- Country of study
- Study design and tool
- Sampling frame
- Sample size
- Inclusion and exclusion criteria, if any
- Type of analysis
- Health condition(s) addressed
- Healthcare services shifted/shared
- Reason for task shifting and/or task sharing
- Cadres of workforces tasks moved from
- Cadres of workforces tasks moved to
- Training provided to enable task shifting and/or task sharing
- Duration of training provided to enable task shifting and/or task sharing
- Guidelines followed to enable task shifting and/or task sharing
- Results and conclusions of the study
- Recommendations of the author(s)

**Stage 5: Collating, summarising and reporting results**

We analysed extracted data thematically [4], using a deductive and inductive hybrid approach [5] to coding data. For the first question “What role has task shifting played in healthcare services delivery since the COVID-19 pandemic?”, we used a top down approach using the WHO Health Systems Framework [6] and checked for impact on each system building block, as well as simultaneous unintended consequences and general acceptability of task shifting. For the second question, “How has the COVID-19 pandemic impacted strategies of task shifting globally?”, we used a bottom up approach to analyse the dataset as per emergent themes. We used the following two coding trees. We discuss our findings in details in the paper.

(A) Role of task shifting in healthcare services delivery since the COVID-19 pandemic

Impact on Health Systems Building Blocks

Service delivery

Health workforce

Health information systems

Access to essential medicines and technologies

Financing

Leadership and governance

Unintended consequences of task shifting

Acceptability of task shifting

(B) Role of COVID-19 on strategies of task shifting

Digital technology enabled capacity building

Modifications in standard operating procedures
