## Supplementary information 3 for "Task shifting healthcare services in the post-COVID world: A scoping review"

| **Study** | **Country of Study** | **Services task shifted** | **HRH delivering services** | **Key findings** |
| --- | --- | --- | --- | --- |
| **COVID-19 testing, surveillance, communication and care management** | | | | |
| Zafar, S. *et al* | Pakistan | Testing, contact tracing and risk communication | Lady Health Workers and Dengue outreach workers | - COVID-19 surveillance hubs were established and CHW were trained briefly before being deployed into communities - As Lady Health Workers and Dengue outreach workers were moved to surveillance, maternal health services and immunisation programs were ignored - Need to improve information systems and TS during emergencies was highlighted |
| Gudza-Mugabe, M. *et al* | Zimbabwe | Testing, contact tracing and reporting | Laboratory technologists | - HRH were trained in antigen-based rapid diagnostic test protocols and testing was decentralised from 1 to over 1000 centres - Access to and uptake of testing increased - Turnaround time of reporting reduced to less than a day - Staff burnouts decreased |
| Honda, C. *et al* | Japan | Contact tracing, reporting and hospital coordination | Part-time PHN | - Acute workloads caused full time PHN fatigue, anxiety and confusion at public health centres - Full time PHN shifted epidemiological surveys, patient hospitalisation coordination, health monitoring of patient contacts and consultation to residents to part-time PHN to prevent burnouts and organisational dysfunction |
| Chidavaenzi, N.Z. *et al* | United States of America | Testing, contact tracing, reporting and quarantining | Public health staff at health department, staff at detention centre and casino (non-healthcare staff) | - Training in rapid antigen-based testing led to 3834 tests for 716 people - Serial testing by non-health staff contributed to reduced case transmissions; only 1 positive case was recorded |
| Raskin, S.E. *et al* | United States of America | Testing, contact tracing, reporting and risk communication via telephone | Dental assistants and hygienists | - Clinic closures and acute demands pulled dental assistants and hygienists into surveillance and community mobilisation - Practices experienced significant call offs from assistants, hygienists and front desk operators, which burdened them further |
| Mohammed, A. *et al* | Nigeria | In-patient COVID care and other essential services | Medical students | - 80.4% students surveyed had good knowledge of COVID-19 - 78.3% students felt at risk of infections, yet 93% of them expressed willingness to assist care provision. Parental disapproval and fear were cited by those unwilling to get involved. - Male respondents were more willing to assist, in comparison to females |
| Taylor, M.K. *et al* | Multi-country (Singapore, Trinidad and Tobago, Iraq, India United States of America, Brazil and more) | Testing, contact tracing, triaging, risk communication and in-patient COVID care | Primary care physicians, medical students | - Primary care physicians tested and triaged COVID-19 patients - Primary care physicians increased awareness about social distancing, symptoms and quarantining - Primary care physicians helped treat cases in intensive care units - Primary care physicians undertook nursing procedures - Final year medical students were involved in in-patient care |
| Eggleton, K. *et al* | New Zealand | Telephone triaging and nursing care | Nurses and practice receptionists (non-healthcare staff) | - Receptionists were upskilled in telephone triaging and they determined whether patients needed GP consultations or referrals - Nurses and receptionist teams ran separate respiratory units to isolate patients with possible symptoms - Routine clinical tasks of COVID-19 were shifted onto nurses - TS increased workloads for nurses and receptionists, but freed GP to attend more patients - TS improved system efficiency and outputs |
| Yoshioka-Maeda, K. *et al* | Japan | Telephone consultations | Office support staff (non-healthcare staff) | - Telephone consultations included queries on prevention measures and patient flow pathways; which did not need PHN involvement - Telephone consultations to be shifted onto low-level staff and office workers through training, creation of script-based manuals and monitoring |
| Yoshioka-Maeda, K | Japan | Telephone consultations, information management, resource management | Office support staff and external company (non-healthcare staff) | - PHN developed a response manual for telephone consultations on COVID-19 and trained teams of office support staff to handle routine queries of local residents - PHN created a web-based system to host patient information management and shifted clerical work to office support staff - PHN shifted management of personal protective equipment to external companies, increasing nursing time and allocative efficiency |
| Helmi, M. *et al* | Indonesia | Intensive care services | GP and medical students | - Intensive care units reported inadequate availability of equipment, service support on call specialists and ineffective TS, leadership and communication among hospital staff - Shifting services from specialists to general practitioners and students needs to be supplemented with training on roles, rights, communication and management skills |
| Sono-Setati, M.E. *et al* | South Africa | Clinical auditing and resource management | Hospital staff (not specific) | - Sub-standard management of COVID-19 cases, medical records and resources and staff anxiety, confusion and stress was reported - Hospitals should expand roles of staff to include TS to address staff shortages and burnouts - Clinical audits should be conducted and reviewed routinely |
| Köppen, J. *et al* | Germany | Emergency procedures for infection control | Emergency paramedics | - Although federal infection control law authorised TS, TS and skill-mixing was not mentioned in state policies directly - Only Saxony‐Anhalt specified the law and provided information supplementing the federal law - Emergency paramedics were allowed to perform certain medical tasks provided they had competencies relevant to standard operating procedures and treatment pathways - Medical directors of services needed to enlist tasks shifted and ensure adequate documentation |
| Faria de Moura Villela, E. *et al* | Brazil | In-patient COVID care and emergency medical services | Physicians, nurses and hospital staff | - TS of services was seen in COVID wards (39.1%), intensive care units (11.6%) and emergency departments (15.9%) - Shifts in roles and corresponding salary cuts coincided with increased anxiety and depression |
| **Mental health screening and therapeutic interventions** | | | | |
| Ortega, A.C. *et al* | Mexico | Psychosocial support, psychological first aid, grief management and palliative mental health services | Primary care physicians, CHW and office staff (non-healthcare staff) | - Non-profit Compañeros En Salud trained non-specialist providers on stress, anxiety and depression screening, identifying nonverbal cues, referral systems, psychological first aid and self-care strategies and provided them with pocket field guides for reference - HRH were trained in contact tracing and their home-visit questionnaires had questions on patient mood, anxiety and suicidal thoughts - Alongside psychological support, they provided mental healthcare for patients who were grieving and patients in need of palliative mental healthcare due to COVID-19 |
| Mukhsam, M.H. *et al* | Malaysia | Psychosocial support | Medical students | - 400 international medical students were tested and quarantined upon return from China - Disinfection and decontamination team performed cleaning upon positive case contact - Quarantine team isolated cases under investigations - Mobile medical and promotion team led surveillance and health education - Students were grouped in tens on online messaging platforms and were supervised by local mandarin-speaking peers, one in each group - Supervisors provided psychosocial support and health education - Supervisors ensured students filled home-monitoring questionnaires |
| **HIV consultation, testing, counselling and treatment services** | | | | |
| Omam, L.A. *et al* | Cameroon | Counselling, testing, patient follow ups, ART initiation and refilling | Primary care physicians, nurses and non-clinical staff | - Primary care-based differentiated service delivery of ART to internally displaced people during COVID-19 was achieved via mobile clinics - In 7 months, 14,623 persons were sensitized and 1,979 were tested, from which 122 tested positive and 33 placed on ART - 28 loss-to-follow up patents were relinked to treatment and 209 consultations were conducted - Mobile clinics were resource-effective in improving access of HIV services in conflict regions, but the model needs economic evaluation |
| Pry, J.M. *et al* | Zambia | ART initiation and refilling, community mobilisation | CHW | - Health ministry COVID-19 mitigation guidelines for HIV recommended dispensing 6 multi-month ART and using TS to communicate and mobilise patients to collect refills early - Adjusted prevalence ratio (4.63; 95% CI 4.45 to 4.82) of early returns to collect ART improved significantly post guideline implementation - Weekly receipt of 6 multi-month dispensation increased from 47.9% before to 73.4% post guideline implementation - Proportion of late visits fell from 18.8% to 15.1% post guideline implementation |
| Abraham, S.A. *et al* | Ghana | Prescribing and dispensing ART | Nurses | - Placing nurses at pharmacies and TS prescribing and dispensing of ART onto them reduced patient clustering and expedited HSD during the pandemic - Staff rosters and roles were changed to complement TS |
| **Others: Sexual and reproductive health; Nutrition; Rheumatoid diseases** | | | | |
| Jacobi, L. *et al* | USA | Provision of contraceptives and dispensing medicines | CHW | - Informants reported that the pandemic has ‘primed’ stakeholders TS and sharing services to and with community health workers at primary care level - TS and sharing provision of contraceptives and dispensing medicines a ‘necessity’ in times of COVID-19 in conflict affected regions |
| Davis, C. *et al* | Singapore | Behaviour contracts for nutrition | GP and nurses | - In-patient services of a tertiary hospital underwent reorganisation of HRH, limiting teams to one physician, nutritionist and speciality nurse - Behaviour contracts for patients and caregivers, charted by psychologists ordinarily, were being drawn up by physicians and nurses during COVID-19 |
| Kuhlmann, E. *et al* | Germany | Care for rheumatoid arthritis and other inflammatory musculoskeletal disorders | GP, rheumatology specialist assistants and other medical assistants | - 67% rheumatologists delegated tasks to rheumatology specialist assistants and other medical assistants - Rheumatologists perceived TS to rheumatology specialist assistants (87%) and GPs (33%) as an efficient approach to address rheumatologist shortages - 81% found cooperation with medical assistants and nurses as good or very good, while cooperation with GPs scored significantly lower |
