## Supplementary information 4 for "Task shifting healthcare services in the post-COVID world: A scoping review"

| **Study** | **Country of study** | **Services task shifted** | **HRH delivering services** | **Key findings** |
| --- | --- | --- | --- | --- |
| **Mental health screening and therapeutic interventions** | | | | |
| Singla, D.R. *et al* | Multi-country (United States of America and Canada) | Screening and therapy for perinatal anxiety and depression | Nurses and midwives | - SUMMIT trial consists of 6-8 weekly sessions on behavioural activation-based therapy offered by trained nurses and midwives in-person or online - Nurses and midwives were trained in 4-day courses and monitored during 8-week internships - Trial is comparing effectiveness of task sharing among telemedicine specialist, telemedicine non-specialist, in-person specialist and in-person non-specialist - Lockdowns led training, supervision, data collection and follow-ups shift online; effectiveness of telemedicine-supported BA is being evaluated |
| Scazufca, M. *et al* | Brazil | Screening, referring and therapy for depression | CHW | - PROACTIVE is a task-shared psychosocial care intervention for depression in adults aged 60 and more, based on psychoeducation and behavioural activation - CHW receive 3 days of training and weekly supervision from clinicians - COVID-19 alerted trial implementation - Recruitment of new participants ceased - Participants who had not completed all sessions were offered 2 telephone sessions substituting in-person programme sessions - 8 month and 12 month follow up was done via telephone - 62·5% participants in the intervention showed recovery from depression compared to 44·0% in the control group (adjusted odds ratio 2·16 [95% CI 1·47–3·18; p<0·0001] |
| Jordans, M.J.D. *et al* | Lebanon | Psychological intervention for children with severe emotional distress | CHW | - Proof-of-concept study evaluated outcomes of competency-driven training in improving facilitator quality - competency-driven training used pre-training and in-training competency assessment outcome scores to adjust dosage of training, focus on competencies and feedback - Lockdowns led training of facilitators move online - Competency-driven training and TS improved training outcomes by 18% without extending class duration |
| Dambi, J. *et al* | Zimbabwe | Screening and therapy for depression | CHW (Grandmothers) | - Physical distancing measures led Friendship Bench to shift onto WhatsApp and telephone to deliver therapy and communicate with patients - Friendship Bench is piloting Inuka, a chat-based application, to further access and enhance help-seeking behavior - A quasi-experimental trial found Iuka to be feasible and demonstrated a decline in common depressive disorders, depression and anxiety and increase in health-related quality of life - Connectivity, app instability, expensive mobile data and power outages were discovered as barriers to scaling up |
| Nirisha, P.L. *et al* | India | Screening and referring patients with alcohol use disorders and mental health for depression | ASHA | - COVID-19 restrictions led trainings adopt hybrid mode - 1-day in-person training method vs 1 day in-person training and digitally-driven 7 online longitudinal training was compared in a trial; screen positives and KAP scores were measured - Online trained ASHAs identified significantly higher number of persons with potential alcohol use disorders [83%; p ≤ 0.001] and common mental disorders [4%; p = 0.018], while in-person trained ASHA identified significantly higher number of those with potential severe mental disorders [61.61%; p ≤ 0.001] - Mean knowledge, attitude and practice score increased from 16.76 to 18.57 (p < 0·01) in ASHA mentored online and from 18.65 to 18.84 (p = 0.76) in in-person trained |
| Philip, S. *et al* | India | Screening and referring patients with substance abuse disorders and depression | Primary care physicians | - 114 primary care doctors were trained, monitored and mentored virtually - Post training case-bases scenario examinations reported 37% improvements in knowledge scores - 80.7% and 47.7% trainees felt confident in identifying mental health issues in patients and their caregivers respectively - 52.6% felt they understand when to refer to higher centres - 60.5% felt confident in prescribing and managing patients with mental health issues - 64.9% respondents felt confident in providing deaddiction services |
| Rodriguez-Cuevas, F. *et a*l | Mexico | Psychosocial support, psychological first aid and grief management | Primary care physicians, CHW, community mental healthcare workers and office staff (non-healthcare staff) | - Non-profit Compañeros En Salud trained non-specialist providers on mental health screening, therapy and referrals - Increase in number of people faced with stress and anxiety due to COVID-19 led intervention to reach COVID patients and families - To mitigate infection transmission risks to CHW, undertook home visits using PPE such as three-layered fabric masks, hand sanitizers (regular visits); N95 or KN95 respirator mask, face shield, isolation gown and gloves (visiting suspected had COVID-19 patients) - They maintained 1.5 metres distance from patients - They met at private ventilated areas other than inside the house, e.g. garden, patio or clinic backyard |
| **HIV consultation, testing, counselling and treatment services** | | | | |
| Coulaud, P. *et al* | Cameroon | Consultation, counselling and ART delivery | Nurses | - Small district hospitals designated to provide HIV services had limited resources and inadequate number of doctors - Hospitals with limited HRH and not practicing task-shifting, reported higher HIV transmission risk and ART stock-outs - Tasks shifting is recommended to maintain ART services delivery as financial resources is being increasingly diverted to pandemic relief |
| Lujintanon, S. *et al* | Thailand | ART initiation | CHW | - Implementation trial is engaging community-based organisations and key population lay providers to start same-day initiation of ART delivery - Lay CHW are trained in counselling, testing, PrEP and ART care - Clients will be offered teleconsultation with physicians and ART home delivery during the COVID-19 pandemic - Success of this trial will help overcome geographical and HRH barriers |
| Roche, S. D. *et al* | Kenya | Consultation, counselling and ART delivery | Hospital staff (not specific) | - One-stop shop model at HIV clinics allowed relocation of drugs, equipment, patient files and PrEP services at one point of patient contact - Task shifting dispensing drugs to lower cadres of care reduced provider movement and patient wait times - However, social distancing protocols caused proximal testing points to shut and few to transform into isolation centres - One-stop shop moved to farther community clinics; patient preferences were unmet - Staff had to travel between centres; wasting productivity |
| **Others: Chronic illnesses including Hypertension, Diabetes; Emergency medical services** | | | | |
| Kamvura, T.T. *et al* | Zimbabwe | Screening of Hypertension and diabetes | Nurses and CHW (grandmothers) | - The Friendship Bench-based intervention is shifting screening of hypertension and diabetes screening onto trained HRH - During lockdowns, grandmothers, nurses and other stakeholders used WhatsApp to disseminate knowledge, advise patients and track referrals |
| Oikonomidi, T. *et al* | France | Consultations and prescribing drugs for chronic illnesses | Digital technology-supported provision of care | - Patients preferred teleconsultations for half of their future consultations - Patients would use online symptom-checkers over contacting doctors for 22.0% of new symptoms - Patients preferred remote monitoring instead of consultations for 52.3% of their treatment adaptations - Prescription renewal and addressing acute or minor complaints were reported as circumstances considered appropriate - Patients expressed that they seek quality assurance and supervision of results by doctors |
| Iwamoto, A. *et al* | Cambodia | Emergency bag-and-mask ventilation, incubator-side tube feeding and temperature measurement | Family caregivers (Fathers and grandmothers) | - Asymptomatic infected family members entering neonatal care units risk transmitting COVID infections to patients and staff - Thermal scanning and frequent hand hygiene were implemented - Implementing asecond line of screening through questions on symptoms and contact tracing is recommended - Risk of transmitting infections necessitates hiring more nursing staff and adequate training and reducing reliance on family caregivers |
